# Automated Evaluation of Antibiotic Prescribing Guideline Concordance in Pediatric Sinusitis Clinical Notes

**DOI:** 10.1101/2024.08.09.24311714

**Authors:** Davy Weissenbacher, Lauren Dutcher, Mickael Boustany, Leigh Cressman, Karen O’Connor, Keith W. Hamilton, Jeffrey Gerber, Robert Grundmeier, Graciela Gonzalez-Hernandez

## Abstract

Ensuring antibiotics are prescribed only when necessary is crucial for maintaining their effectiveness and is a key focus of public health initiatives worldwide. In cases of sinusitis, among the most common reasons for antibiotic prescriptions in children, health-care providers must distinguish between bacterial and viral causes based on clinical signs and symptoms. However, due to the overlap between symptoms of acute sinusitis and viral upper respiratory infections, antibiotics are often over-prescribed.

**Objectives:** Currently, there are no electronic health record (EHR)-based methods, such as lab tests or ICD-10 codes, to retroactively assess the appropriateness of these prescriptions, making manual chart reviews the only available method for evaluation, which is time-intensive and not feasible at a large scale. In this study, we propose using natural language processing to automate this assessment.

**Methods:** We developed, trained, and evaluated generative models to classify the appropriateness of antibiotic prescriptions in 300 clinical notes from pediatric patients with sinusitis seen at a primary care practice in the Children’s Hospital of Philadelphia network. We utilized standard prompt engineering techniques, including few-shot learning and chain-of-thought prompting, to refine an initial prompt. Additionally, we employed Parameter-Efficient Fine-Tuning to train a medium-sized generative model Llama 3 70B-instruct.

**Results:** While parameter-efficient fine-tuning did not enhance performance, the combination of few-shot learning and chain-of-thought prompting proved beneficial. Our best results were achieved using the largest generative model publicly available to date, the Llama 3.1 405B-instruct. On our test set, the model correctly identified 91.4% of the 35 notes where antibiotic prescription was appropriate and 71.4% of the 14 notes where it was not appropriate. However, notes that were insufficiently, vaguely, or ambiguously documented by physicians posed a challenge to our model, as none evaluation sets were accurately classified.

**Conclusion:** Our generative model demonstrated strong performance in the challenging task of chart review. This level of performance may be sufficient for deploying the model within the EHR, where it can assist physicians in real-time to prescribe antibiotics in concordance with the guidelines, or for monitoring antibiotic stewardship on a large scale.

## 1. Introduction

Antibiotic stewardship programs (ASPs) aim to optimize the use of antibiotics for specific conditions and to combat the growing threat of antimicrobial resistance.^1^ Inappropriate prescribing of antibiotics not only contributes to a global health crisis but also exposes patients, particularly pediatric patients, to unnecessary side effects and disrupts their healthy microbiota.^2^ Ensuring that antibiotics are prescribed adequately—only when necessary and with the correct dosage and duration—is essential for maintaining their efficacy and is a key focus in public health and research efforts at national and international levels.

Most antibiotic prescribing takes place in the ambulatory setting, and approximately 30% of all outpatient antibiotic prescriptions are unnecessary; a majority of unnecessary outpatient prescribing is for acute upper respiratory tract infections.^3,4^ In particular, sinusitis which is among the most common reasons for ambulatory antibiotic prescribing in children.^3^ The symptoms of acute sinusitis often overlap significantly with those of uncomplicated viral upper respiratory tract infections. As a result, antibiotics are often over-prescribed for sinusitis, despite guidelines recommending more conservative use.^5,6^

The Centers for Disease Control and Prevention (CDC) Core Elements of Outpatient Antibiotic Stewardship recommend tracking and reporting ambulatory antibiotic prescribing.^7^ Some metrics using data from the electronic health record (EHR) have been developed in order to measure unnecessary and guideline-discordant prescribing, which can be used in feedback for clinicians and practices and in assessing the impact of stewardship programs on prescribing.^8,9^ However, determination of appropriateness using an electronically-based metric is more challenging for some conditions, such as sinusitis. Healthcare providers must distinguish bacterial from viral sinusitis based on clinical signs and symptoms alone, and antibiotic prescribing is only considered guideline-concordant for bacterial sinusitis. As such, there are no lab tests or ICD-10 codes that can be used to retroactively measure prescribing appropriateness in the absence of time-intensive manual chart review.

Traditionally, audits of patient charts for assessing antibiotic prescription guideline concordance have relied on manual review by infectious disease specialists.^9–11^ While this approach has elicited important findings for antibiotic stewardship, including discordant antibiotic prescribing and selection, there are limitations to manual review. Retrospective manual review of charts is labor intensive and time consuming, therefore only small samples of charts can be reviewed and the selection of these charts, if based on ICD codes, may be less than optimal as these codes may be incorrectly assigned or missing.^11^

To date, limited research has been undertaken to automate chart review for antibiotic appropriateness assessment. Several studies have created classification models to assess appropriate antibiotic prescribing by linking patient diagnoses to tier-based rules where the antibiotic prescription is always, sometimes, or never appropriate depending on the diagnosis.^12–14^ While these classification models performed reasonably well, they do not take into account specific patient variables that may influence prescribing decisions. Our study is the first attempt to utilize advanced AI methods to identify relevant patient information in the free text of clinical notes and automatically apply the logical criteria of prescribing guidelines to assess antibiotic prescribing appropriateness.

This paper explores the significance of antibiotic stewardship for pediatric sinusitis and presents a generative system, utilizing a Large Language Model (LLM) approach, to automate the analysis of unstructured notes from the CHOP primary care practices to determine justified vs unjustified prescription of antibiotics given a case presentation, seeking to enable a large-scale study that aims to improve prescribing practices.

## 2. Materials and Methods

We represented the task of evaluating the guideline concordance of antibiotic prescribing in clinical notes as a decision task. That is, given a note in which a patient was diagnosed with sinusitis and prescribed antibiotics, our system should predict whether the prescription was 1) appropriate, 2) not appropriate, or 3) insufficient or ambiguous, in cases where the note does not contain enough information to assess the appropriateness of the prescription.

### 2.1. Data collection

We identified all pediatric (younger than 18) clinical encounter notes by ICD-10 code from outpatient billed encounters at one of 32 primary care practices in the Children’s Hospital of Philadelphia (CHOP) network from July 1, 2017 through June 30, 2021 using the following criteria: 1) visits with either a J01 (acute sinusitis) or J32 (chronic sinusitis) code and 2) a prescription of an oral antibiotic (excluding antibiotics that would never be prescribed for sinusitis). The following patients were excluded: 1) patients with a confounding chronic medical condition identified by an ICD-10 code;^15^ 2) patients with an ICD-10 code for another infection that would warrant an antibiotic prescription at the same visit.

A total of 10,311 patients met the inclusion criteria 6,377 (61.9%) for acute sinusitis, and 3,934 (38.2%) for chronic sinusitis, seen by 310 providers. The median number of encounters per provider was 12 (3 – 48).

To develop, train, and evaluate our classifier, we selected 300 encounter notes at random. Our intent was to reflect the natural distribution of the notes where the system will be deployed, so we did not oversample or undersample any specific group or provider. This resulted in 190 (63.3 %) encounter notes for acute sinusitis and 110 (36.7%) for chronic sinusitis, seen by 132 providers. The median number of encounters per provider was 50 (21.5 – 92).

We split our annotated dataset into three sets, the first two of which were selected from 80 percent of the providers: a training set with 200 notes (117 notes with appropriate prescriptions, 69 not appropriate, and 14 with insufficient or ambiguous documentation), a development set with 50 notes (32 appropriate, 16 not appropriate and 2 insufficient). For the third set (the test set), we selected 50 notes from the remaining 20% of the providers (35 appropriate, 14 not appropriate, 1 insufficient), in order to be able to test the system on how it adapts to notes from new (unseen) providers.

### 2.2. Annotation

We derived a set of criteria by adapting the recommendations of two clinical practice guidelines^16,17^ to define the appropriateness of antibiotic prescribing to the patients we selected. Table 1 summarizes our criteria. If a patient met at least one criterion, our annotators labeled the note as appropriate. If there was clear evidence in the note that none of the criteria were met, the annotators labeled the note inappropriate; otherwise, if it was not possible for the annotator to decide if the criteria were met or not in an note, the note was labeled insufficient. Such cases usually include incomplete documentation, or ambiguous or vague descriptions. The phrase “*patient had congestion for over a week* “ is an example of an ambiguous description. If the congestion lasted for 8 or 9 days, criterion 1 in Table 1 would not be met and this would labeled ‘not justified’. However, if the symptom lasted 10 days or longer, then criterion 1 would be satisfied and this would be labeled ‘justified’. The phrase “*Fever x 3 days*” is an example of incomplete documentation because it does not specify the exact temperature. Note that our definition focuses solely on the act of prescribing antibiotics and excludes considerations related to the appropriateness of the specific antibiotic prescribed, as well as its dosage and duration.

**Table 1:** Clinical guidelines used to assess the appropriateness of an antibiotic prescription for patients diagnosed with sinusitis. If the clinical note provided sufficient evidence to meet at least one of the three established criteria, the prescription was annotated as appropriate.

| Antibiotics appropriateness |
| --- |
| 1. Persistent illness: nasal discharge (of any quality), daytime cough, or sinus pain/pressure lasting for $\geq 10$ days without improvement |
| 2. Severe onset, i.e., concurrent fever (temperature $\geq 39^{\circ}\text{C}/102.2^{\circ}\text{F}$) and purulent nasal discharge or sinus pain/pressure for at least 3 consecutive days |
| 3. Worsening course, i.e., worsening or new onset of nasal discharge, daytime cough, sinus pain/pressure, or fever after initial improvement |

One pediatric physician annotated the 300 notes of our corpus as *appropriate, inappropriate*, or *insufficient*. A second pediatric physician is currently annotating 50 notes of our corpus to compute the inter-annotator agreement. To guide this assessment, an annotation guide was developed, using input from a primary care pediatrician, two infectious diseases specialists, and one pediatric infectious diseases specialist. This annotation guide was developed iteratively using practice notes from the same practices with the goal of improving reproducibility as much as possible.

### 2.3. Generative models

To successfully complete this task, our classifier must extract the symptoms of interest from the clinical notes -congestion, cough, sinus pain/discomfort, and fever-assess their severity, and understand their time or progression patterns. The classifier then needs to perform logical validation by verifying whether the extracted data meets the logical rules defined by our criteria Table 1. Information is often dispersed across various sections of a note, requiring the reader to infer details that may be implicitly documented. For instance, the reader may need to resolve deictic references like “*cough started last Wednesday* “ by determining the number of days from the onset of the cough to the date of the visit always noted at the beginning of the note. This task poses a challenge for conventional natural language processing systems. These systems often employ a pipeline approach, resolving each task independently. However, this approach can propagate errors from earlier stages and introduce new ones in later stages, which can limit the system’s overall performance.^18,19^

As an alternative to conventional Natural Language Processing (NLP) systems, in this study, we propose using state-of-the-art generative systems powered by large language models (LLMs). In recent years, generative systems have become the leading approach in NLP as evidenced by the widespread success of chatGPT.^20^ Generative systems feature interfaces that allow users to submit prompts in natural language, an intuitive interface to perform a task.^21^ These prompts typically include an instruction specifying the desired action, along with optional data needed to perform the task. Generative systems leverage semi-supervised training to transfer general knowledge acquired from extensive text corpora, enabling them to generate appropriate responses and execute instructions for tasks they were not explicitly trained on. This eliminates the need to retrain the system for each specific task, a requirement often necessary in conventional NLP systems.

One key factor crucial in the development of generative systems was easy access to massive amounts of text such as the Common Crawl project,^22^ which at the time of writing counts 3.35 billion pages from the internet that can be freely downloaded. These texts provide real-world examples to pretrain language models with semi-supervision. Pretraining algorithms may differ but they rely on the same general idea: given a sequence of written text and a set of words masked in the sequence, the model must predict the words hidden. Erroneous predictions trigger changes to the weights of the neural networks encoding the language model to improve the likelihood of predicting the hidden words at the next iteration. For example, in the sequence *During an episode of pain, an EKG reveals [***]*, a clinically trained large language model will guess ‘ST depressions’ as it has encoded weights that memorized that *ST depressions* (the hidden words) is a common sign associated with chest pain because they were seen co-occurring multiple times in other texts. The exact mechanisms by which neural networks capture such knowledge are still unclear, but emerging evidence indicates that, from these massive correlations of phrases, language models learn structured and grounded representations of objects existing in our world and how they interact, allowing them to make generalizations and provide reasonable answers to various questions even when tackling unseen tasks. For instance, Gurnee & Tegmark^23^ demonstrated the existence of linear representations of space and time in the Llama 2 family of generative models.^24^ They probed the network and show the existence of a representation explicitly encoding the dates of death of public figures which can be called upon by the models to successfully answer “who died before whom” questions, among others, without having seen the answers in any written texts the models were trained on.

More powerful processors and better training algorithms work in concert to improve the training speed of neural networks, allowing the scaling up of the language models from millions^25^ to hundreds of billions of parameters.^26^ Faster Graphics Processing Units (GPUs) have been embraced by the AI/NLP community to accelerate matrix calculations which is fundamental for training neural networks. The transformer architecture, which allows parallel computing of their components,^27^ is currently the standard for NLP.^28^ It stacks multiple layers of multi-head self-attention to encode efficiently the meaning of sentences/paragraphs into a multidimensional space, using similarity between phrases seen co-occurring during the pretraining of the language model. The core mechanism of self-attention helps a neural network to learn various syntactic, semantic, and pragmatic relations between the phrases composing the sentences, computed in parallel. These relations are used as features by the generative systems to perform the task at hand. The increase in generative model sizes enhances the performance of the models on known tasks but also unlocks new capabilities that only the biggest models can accomplish.^29^ Models sufficiently large develop the *chain of thought* followed by humans when reasoning.^30^ That is, the model mimics the logical steps followed by humans when solving a problem and, by learning how to explain their reasoning, they improve their performance on the task.

### 2.4. Classification with Generative systems

We performed our classification using generative systems from the Llama 3 family,^26^ which is one of the largest freely available sets of models offering competitive performance compared to proprietary alternatives. We progressively refined an initial simple prompt by following few-shot learning and chain-of-thought techniques to enhance the models’ performance on our task. Additionally, we fine-tuned a Llama-3-70B-Instruct model using a parameter-efficient fine-tuning (PEFT) approach, namely LoRA,^31^ to specialize the model for our specific task.

#### Initial prompt

Figure 1 outlines the various components of the prompt we designed to instruct our model on how to classify the appropriateness of antibiotic prescription in clinical notes. We began our experiments with an initial straightforward prompt that defined the role the generative model should assume, followed by a brief paragraph specifying the instructions for the task. This paragraph included the following key components:

**Fig. 1:**
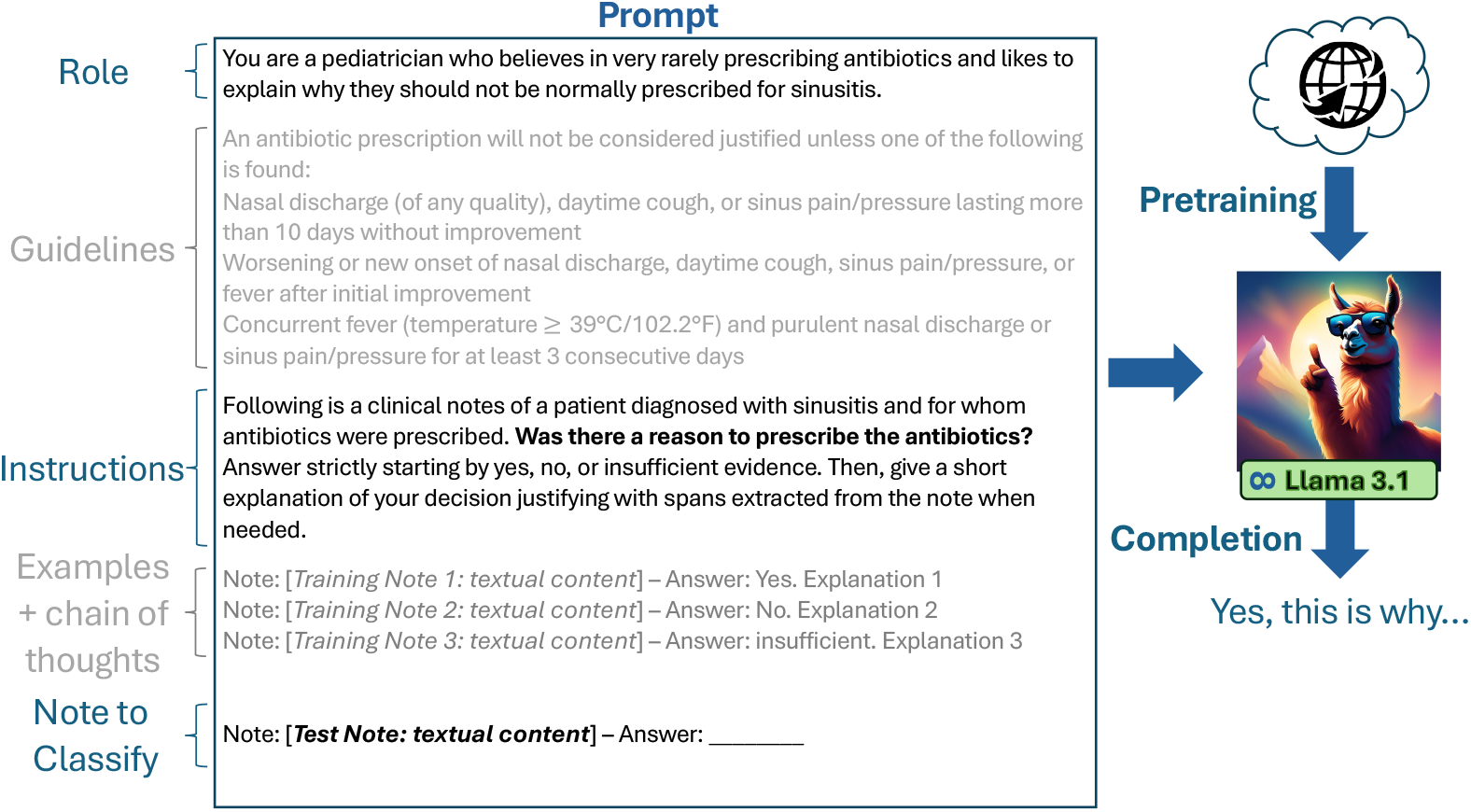
Iterative construction of a prompt to classify antibiotic prescription appropriateness using a Llama 3 generative model.

1. The role specifying the function the model should adopt when generating response, in our case a pediatrician.
2. The context which describes the notes; specifically, the input note is a clinical note of a patient diagnosed with sinusitis who received antibiotics.
3. The question the model should answer.
4. The format in which we wanted the model to present its response.
5. The text of the note to be classified
6. The keyword *Answer:* to initiate the model’s completion according to our specified format.

The authors, during an interactive session, tried multiple initial prompts and evaluated the Llama 3 70B-instruct model’s results on the development set. At the end of the interactive session, we selected the initial prompt illustrated in Figure 1.

We first extended this initial prompt by inserting the clinical guidelines that our annotators followed when labeling the notes in our corpus. The generative models that are publicly available were pretrained on large corpora from the internet, which contain few, if any, professional medical documents^29,32^ and may not have encountered or memorized our specific guidelines. By including these guidelines directly in the prompt, we ensured that the model had direct access to the criteria defining the task. The guidelines in Table 1 were written for medical professionals. To make them more accessible and easier for our generative model to interpret, *LD*, Assistant Professor in Medicine, simplified the language used in the guidelines.

#### Few-shot learning

Much like humans, generative models can benefit from seeing a few examples before attempting a task, a concept known as few-shot learning. We implemented this approach by including the text of three notes from our training set in the prompt, each accompanied by their appropriateness labels. Although complex conditions could be used to select these examples -such as choosing notes with close semantic similarity to the one being classified or those that annotators found challenging^33^-we opted to select the examples randomly. We chose this approach for simplicity and left the exploration of more sophisticated selection strategies for future work.

#### Chain-of-thought prompting

Together with few-shot learning, we also employed chain-of-thought prompting. After each label of our training examples, we included a brief explanation of the label, along with the relevant quotes that demonstrated the extracted span from the example note supporting the explanation. *LD* provided these explanations, highlighting which criteria from Table 1 were met, missing, or challenging to verify based solely on the note’s text. Requesting explanations along with quotes forces the model to ground its responses within the text of the notes, thereby reducing hallucinations. Despite the large context window of 8,192 tokens, the Llama-3-70B-Instruct model still has a limited prompt capacity, which restricted us to including no more than three example notes. In a supplementary experiment, to include additional examples, we did not input the entire text of the training notes. Instead, we only incorporated the sentences containing the relevant quoted phrases.

We hypothesized that chain-of-thought reasoning is an important component for improving the performance of a generative model, and conducted additional experiments by reformulating our initial prompt and its components. While the description of the model’s role remained unchanged, we revised the context and question to predispose the model to answer ‘not appropriate’ by default unless it identified evidence in the notes that satisfied a criterion from our guidelines. We also introduced simple definitions for *’fever,’ ‘nasal discharge,’ and ‘nasal congestion’* before presenting the guidelines, and we rephrased the guidelines as a set of eight conditional rules. Additionally, we revised the explanations for all ten examples in the prompt to explicitly indicate which rules were met or unmet (Line 9 in Table 2) (we provide the exact prompt used in these experiments in the Appendix A).

**Table 2:**
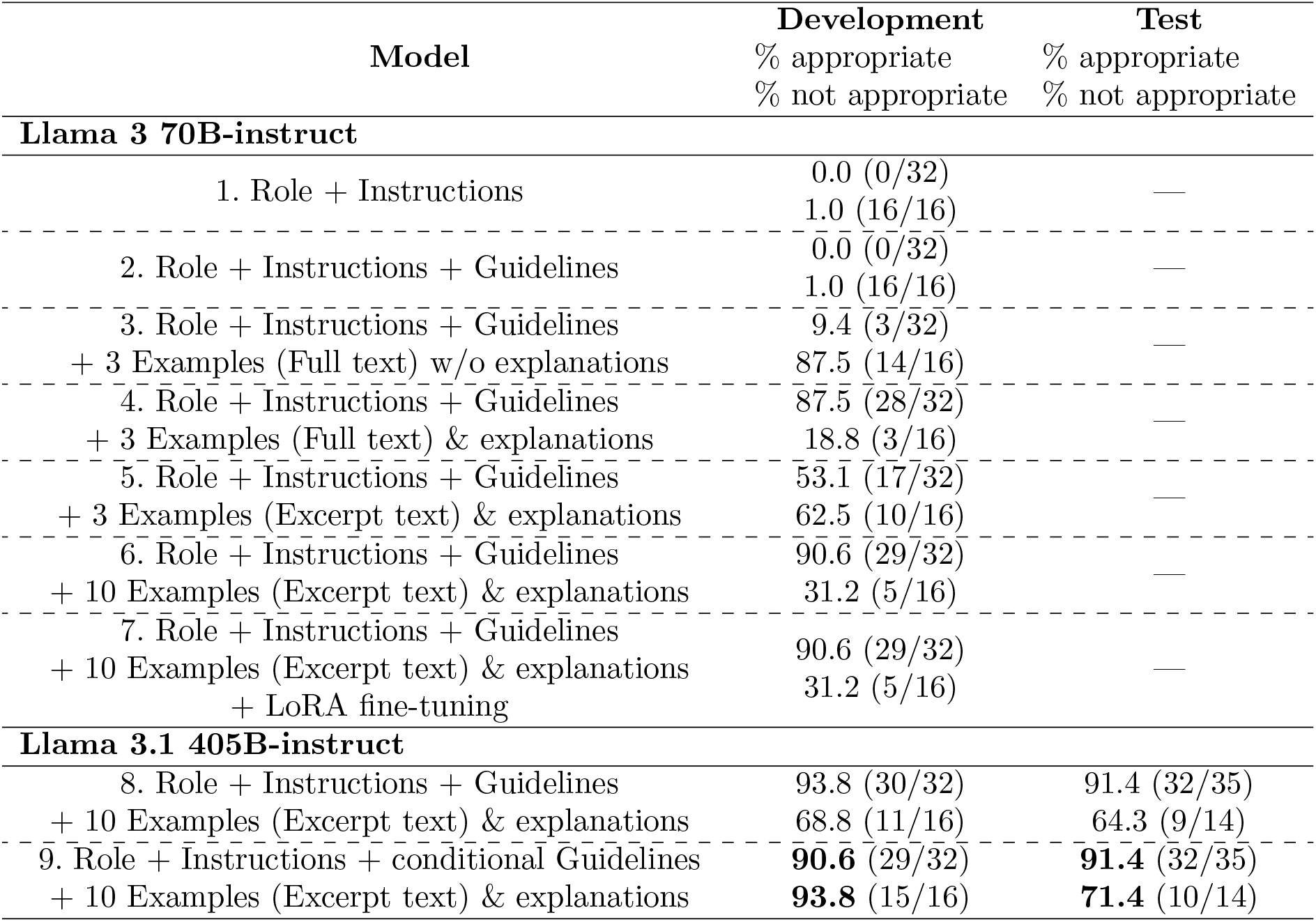
Classification results on the development and test sets for various prompts and large language models. We omitted the percentages of notes labeled as insufficient because none of the models were able to retrieve any in this category.

#### Parameter Efficient Fine-tuning with LoRA

Although generative models achieve state-of-the-art performance on general NLP tasks, they may benefit from being fine-tuned to perform more specific and challenging tasks. A standard method for training generative models is full fine-tuning, a supervised training process. In this process, the model is presented with instructions and corresponding data required to perform a task. It generates a response, which is then automatically compared to the expected gold-standard answer from the training examples. If the model generates the expected answer, no adjustments are made. However, if it deviates, all weights of its underlying neural network are updated to increase the likelihood of generating the correct response. While fine-tuning can enhance the model’s performance on specific tasks, updating all weights of a very large neural network is computationally intensive and requires a significant number of expensive GPUs, which were not available for our experiments. LoRA is a heuristic proposed by Yu et al.^31^ to update only a small portion of the weights in the neural network. We trained the Llama 3 70B-instruct model using the implementation of LoRA from the litGPT^34^ with the rank set to 32 for 20 epochs. We retained the checkpoint that achieved the best score on our development set as the trained model.

#### Evaluation

We evaluated the performance of the Llama 3 70B-instruct model using our initial prompt on the development set, then assessed its performance as we sequentially added each component designed to enhance the prompt—namely, guidelines, a few examples, explanations for the labels, and finally, fine-tuning on our training corpus. Recognizing that the performance of generative systems typically improves with larger underlying language models, we also tested the best-performing setup with the Llama 3 70B-instruct model on the recently released Llama 3.1 405B-instruct quantized (int4) model. ^a^ We conducted all experiments with the Llama 3 70B-instruct model with a temperature set to 0.001, top-p to 0.01, and top-k to 1, ensuring deterministic responses from the system by consistently selecting the most likely token when generating its answers. For our evaluation, we conducted all experiments with the Llama 3.1 405B-instruct model with a temperature set to 0.6, top-p to 0.9, and top-k to 50, allowing more variety in its responses. Since there were very few notes labeled as insufficient in our gold standard, most errors involved the model confusing notes with appropriate (guideline-concordant) prescriptions with those that were not appropriate (not guideline-concordant) and vice versa. Therefore, we chose to report all results by only providing the percentage of notes in each class that were correctly labeled by the generative model and did not report the more standard F1-scores.

## 3. Results and Discussion

We present our results in Table 2, highlighting best performance on the test set. The system correctly identified 10 out of 14 (71.4%) notes labeled as not appropriate and 32 out of 35 (91.4%) notes labeled as appropriate (line 9.). This performance was achieved by providing the model with instructions and logical guidelines to perform the task, without extensive training on the training set. The classifier demonstrated remarkable correctness on a complex task typically performed by trained physicians when only given a few examples and clear explanations indicating whether the rules of the guidelines were satisfied or not.

The table shows that all modifications made to the initial prompt (line 1.) led to incremental improvements in the model’s classification performance. The table offers several interesting insights. Firstly, it is surprising that truncating the text of the example notes did not lead to a performance drop (line 4. *vs*. line 5.). This suggests that most of the text in a note is not utilized by the model for understanding the examples and can be omitted without losing essential information. Secondly, it is worth noting that LoRA, the parameter-efficient technique we employed on our training set, did not enhance the model’s performance (line 6 vs. line 7).This unexpected result requires additional experiments for further explanation. Thirdly, our findings align with recent trends in the Natural Language Processing community, which indicate that generative models based on larger language models perform better than their smaller counterparts. This is evident in Table 2, where the Llama 3.1 405B-instruct model, at the time of writing, the largest model freely available to the community, outperformed the Llama 3 70B-instruct model. Lastly, since the model was not specifically trained on our training set, it was not biased toward recognizing the style of certain providers over others. It demonstrated robustness to variations in providers’ styles and achieved comparable performance on the test set as it did on the development set.

### 3.1. Error analysis

We analyzed the errors made by the best classifier, the Llama 3.1 405B-instruct model (line 8 in Table 2), on the examples in the test set. The model misclassified a total of 8 notes. The most frequent errors were False Positives (FPs), where the notes were labeled as inappropriate for antibiotic prescription, but the classifier predicted them as appropriate. There were 5 such misclassified notes. Upon re-examining the notes, *LD* reviewed the explanations provided by the model and determined whether they were valid. It was found that 2 FPs occurred in notes that could have been labeled as insufficient due to ambiguous temperature documentation. For the remaining 3 FPs, *LD* confirmed the errors made by the system. One error resulted from the incorrect resolution of a deictic time reference; another from a misinterpretation of the term ‘worsening’ (in the phrase ‘acutely worsening symptoms overnight’, where ‘worsening’ refers to an increase in the severity of symptoms, not the progression pattern where the patient initially feels sick, then slightly better, and then worse); and the final FP was due to the system’s hallucination, incorrectly stating that a temperature of 102°F is higher than 102.2°F. The model had more success classifying the notes in which antibiotics were prescribed appropriately. There were only 3 False Negatives (FNs), as these notes clearly mentioned the onset and duration of symptoms. One FN occurred due to the under-specification of the definition of fever in criterion 3 in Table 1; unlike criterion 2, the exact temperature defining a fever is not specified. As a result, there was a disagreement between the annotator and the system regarding the resolution of this criterion in the note. The last two FNs were made on notes that were ambiguous and could have been labeled insufficient.

Finally, the note labeled as insufficient was misclassified by the model. Although the model correctly extracted the phrase “*congestion for over a week* “ vaguely documenting the timing of symptom onset, it failed to recognize the ambiguity and opted for the shorter duration, leading to the wrong conclusion that the antibiotic prescription was not appropriate.

#### 3.2. Limitations and future work

The largest model, Llama 3.1 405B-instruct, demonstrated impressive performance on our task. It was able to follow the logic of our guidelines and provide reasonable explanations for its decisions without explicit training. Although the task is challenging, it only requires the system to identify four common symptoms, assess their severity, and understand their duration or progression patterns. As evidenced by our performance with general generative models, the necessary knowledge to perform the task was available in their training data from the internet. However, most clinical NLP tasks will require specialized knowledge available only in clinical notes and ontologies. Researchers will need to continue pretraining or fine-tuning these models to integrate this domain-specific knowledge. As the size of generative models continues to grow, these training tasks become increasingly challenging for standard institutions such as hospitals and universities, which may lack the necessary hardware for the required computations.^35^

We acknowledge that a sample size of 50 notes is relatively small for evaluating our generative model. However, since our system does not benefit from LoRA training, we will evaluate its performance on the entire training set, providing a more comprehensive assessment. Ambiguous and vague documentation in the notes remains a challenge for our best model, as most errors occur on these notes. With larger language models now supporting input prompts of up to 16,000 tokens, we plan to include more examples of vague and ambiguous notes, along with explanations, to help the model recognize and classify these cases appropriately. Despite forcing the models to justify their decisions and anchor their answers within the input texts, we still found instances of hallucination. Integrating ‘debates’ among several generative LLM-based models has been proposed as an effective solution to detect and reduce hallucinations.^36,37^ Our approach could easily be extended from a single generative model performing classification to a deliberative panel finding consensus for each debated note. We leave the deployment and evaluation of this approach to future work.

## 4. Conclusion

To address the challenge of over-prescribing antibiotics for sinusitis in children, this study proposes using natural language processing to automate the assessment of prescription appropriateness, overcoming the limitations of time-consuming manual chart reviews. We developed, trained, and evaluated generative models to classify the appropriateness of antibiotic prescriptions in 300 clinical notes from pediatric patients with sinusitis at the Children’s Hospital of Philadelphia primary care network. Although Parameter-Efficient Fine-Tuning did not improve performance, the combination of few-shot learning and chain-of-thought prompting proved beneficial. Our best results were achieved using the largest generative model available at the time, the Llama 3.1 405B-instruct. On our test set, the model correctly identified 91.4% of the 35 notes where the antibiotic prescription was appropriate and 71.4% of the 14 notes where it was not. Without extensive training, our generative model demonstrated strong performance in this complex task, suggesting it could be effectively deployed within the EHR to assist physicians in real-time to prevent over-prescribing as well as in monitoring antibiotic prescribing on a large scale.

## Data Availability

The data used in this study contain protected health information (PHI) and are not publicly available due to privacy and confidentiality regulations.

## Acknowledgements

The research reported in this publication was supported by a Centers for Disease Control and Prevention (CDC) Cooperative Agreement Funding Opportunity Announcement (FOA) U54 CK000610, Epicenters for the Prevention of Healthcare-Associated Infections.

## Appendix A

### Prompt with conditional guidelines

#### Role

You are a pediatrician who believes in very rarely prescribing antibiotics and likes to explain why they should not be normally prescribed for sinusitis.

#### Definitions

A patient has fever if the patient has a body temperature above 102.2°F or 39°C. Nasal discharge quality can be a. clear and watery, b. thick and white, c. yellow, green, brown or grey, d. bloody, e. foul-smelling, f. thick and stringy, g. purulent. If a patient has nasal passages congested, then the patient has nasal discharges.

#### Conditional guidelines

As a pediatrician you would consider that the prescription of an antibiotic was not justified unless one or more of the following rule is satisfied:

Rule 1. If the patient had nasal discharge of any quality for 10 or more days without improvement, then the prescription of the antibiotics was justified.

Rule 2. If the patient was coughing during the day for 10 or more days without improvement, then the prescription of the antibiotics was justified.

Rule 3. If the patient experienced discomfort or pain in the areas around the sinus for 10 or more days without improvement, then the prescription of the antibiotics was justified.

Rule 4. If the patient had nasal discharge of any quality, was recovering, but then experienced an increase of its severity or its reappearance, then the prescription of the antibiotics was justified.

Rule 5. If the patient was coughing during the day, was recovering, but then experienced an increase of its severity or its reappearance, then the prescription of the antibiotics was justified. Rule 6. If the patient experienced discomfort or pain in the areas around the sinus, was recovering, but then experienced an increase in the pain/discomfort severity or its reappearance, then the prescription of the antibiotics was justified.

Rule 7. If the patient had fever and, in the same time, had purulent nasal discharge for 3 or more consecutive days, then the prescription of the antibiotics was justified.

Rule 8. If the patient had fever and, in the same time, experienced discomfort or pain in the areas around the sinus, for 3 or more consecutive days, then the prescription of the antibiotics was justified.

#### Instructions

Following is are clinical notes of patients diagnosed with sinusitis and for whom antibiotics were prescribed. By default, assume that the prescription of antibiotics was not appropriate unless you find evidence in the note indicating that at least one of the preceding rules was satisfied. Answer strictly starting by Yes, No Insufficient. Then, give a short explanation of your decision justifying with spans extracted from the note when needed.

#### Examples - (note excerpt, answer, explanation, quotes)

Note: ‘*note 1 excerpt* ‘ Answer: Yes. Explanation: The patient was coughing for two weeks which is more than 10 days therefore the prescription of antibiotics was justified (Rule 2. is satisfied) Quote: “cough x 2 weeks”

Note: ‘*note 2 excerpt* ‘ Answer: No. Explanation: The patient had nasal passages congested for only 5 days which is less than 10 days (Rule 1. is not satisfied). The patient experienced pain in the areas around the sinus pain but no duration was specified (Rule 3. is not satisfied). Quote: “Thick yellow green congestion for about 5 days”

Note: ‘*Note 3 excerpt* ‘ Answer: Insufficient. Explanation: The patient was coughing during the day between 7 to 10 days. This duration is vague. If the patient was coughing for less than 10 days then the prescription of the antibiotics was not justified (Rule 2. is not satisfied), on the contrary, if the patient was coughing for 10 days then the prescription of the antibiotics was justified (Rule 2. is satisfied) Quote: “Cough? 7-10 days”

#### Note to classify

Note: ‘*textual content of the note to classify*’ Answer:

## Footnotes

a The Llama 3.1 405B was released on July 23, just nine days before the submission deadline for this publication. Consequently, we were unable to rerun all experiments with this model, as it takes approximately 30 minutes to classify a single note.

